# Safety of MVA-BN in Healthcare Personnel, Democratic Republic of the Congo

**DOI:** 10.1101/2025.06.11.25328898

**Authors:** Faisal S. Minhaj, Anna Mandra, Béatrice U. Nguete, Stephanie Tran, Jordan L. Kennedy, Benjamin Monroe, Christine M. Hughes, Taina Joseph, Marissa K. Person, Michael B. Townsend, Panyampalli S. Satheshkumar, Toutou Likafi, Gaston Kokola, Joelle Kabamba, Mary G. Reynolds, Agam K. Rao, Dorah Kasongo, Patricia A. Yu, Yon Yu, Robert Shongo Lushima, Didine Kaba, Brett Petersen, Andrea M. McCollum

**Affiliations:** Division of High Consequence Pathogens and Pathology, Centers for Disease Control and Prevention, Atlanta, GA, USA; Kinshasa School of Public Health, Democratic Republic of the Congo; Ministry of Health, Kinshasa, Democratic Republic of the Congo; Centers for Disease Control and Prevention, Kinshasa, Democratic Republic of the Congo; Office of Readiness and Response

## Abstract

**Background:** Modified vaccinia Ankara-Bavarian Nordic (MVA-BN) is a third generation, replication-deficient, smallpox and mpox vaccine prepared in liquid and lyophilized formulations. The Democratic Republic of the Congo reports the highest number of cases of clade I mpox annually.

**Methods:** During the summer of 2017 and 2019, 1,600 healthcare personnel were enrolled in an investigational prospective cohort study to evaluate safety of the 2-dose MVA-BN vaccine series; 1000 were administered liquid and 600 were administered reconstituted lyophilized MVA-BN. Vaccine doses were given on study days 0 and 28 and presence or absence of adverse events were evaluated at routine follow-up appointments for up to two years.

**Results:** There were 49% of liquid and 54% of lyophilized formulation participants who experienced at least one adverse event within 7 days. Eighteen serious adverse events, including seventeen deaths and one stillbirth, occurred within the 2-year study period; safety monitors deemed none were causally associated with vaccine administration. Fourteen pregnancies occurred within one month of vaccination, 13 gave birth to healthy infants.

**Conclusions:** This study adds to the growing body of literature on the safety of MVA-BN in different populations. The data from this Congolese cohort demonstrate good safety outcomes for up to two years with both liquid and lyophilized vaccine formulations. As MVA-BN vaccination campaigns are launched in endemic and surrounding countries in late 2024 as part of clade I mpox outbreak response efforts, the data from this study supporting safety of MVA-BN in an African population are crucial to build vaccine confidence.

Trial registry number: NCT02977715

## Introduction

Mpox is caused by monkeypox virus which has two clades: clade I is found in Central Africa and is historically associated with more severe disease; clade II is found in West Africa and spread globally starting in 2022. The Democratic Republic of the Congo (DRC) is endemic for clade I MPXV and continuously reports the majority of African mpox cases, predominantly occurring in forested areas of the country. Since 2023, DRC has experienced an unusual outbreak of suspected clade I MPXV affecting provinces that previously had not reported human cases; the outbreak has further spread to neighboring countries and via travel to other continents. Healthcare personnel in DRC are known to be at least 3 times higher risk for infection than the general population.^1^ This may be due to differences in healthcare personnel training, limited resources, and lack of infection prevention and control measures.

Modified vaccinia Ankara-Bavarian Nordic (MVA-BN) or JYNNEOS^®^ is a live, third generation vaccine licensed for smallpox and mpox prevention in the United States and does not replicate in mammalian cells.^2^ MVA-BN has been studied in special populations in high income countries including persons living with HIV^3^, atopic dermatitis^4^, and hematopoietic stem cell transplant recipients.^5^ Vaccine use in these populations was safe and well tolerated in clinical studies. Additional studies evaluated cardiac safety due to other live, replicating orthopoxvirus vaccines known association with myopericarditis.^6^ MVA-BN was extensively used in high income countries throughout the global mpox outbreak that started in 2022 and passive surveillance through the Vaccine Adverse Reporting System and the Vaccine Safety Datalink supported the safety of MVA-BN in broad populations in the United States, including patients with HIV.^7,8^ MVA-BN is manufactured as both a liquid and as a freeze-dried frozen formulation, however only the liquid form has been used outside of clinical trials including during the clade II global outbreak. More recently it is being used in Africa in 2024-2025 to combat the clade I outbreak. The liquid formulation is easier to use once thawed as it can be drawn up directly from the vial for administration. The lyophilized formulation must be reconstituted, adding an additional preparation step, however, it has the advantage of improved stability at refrigerated temperatures (i.e., 2-8°C).

In 2017, we initiated a study to evaluate the safety and immunogenicity of liquid and lyophilized formulations of MVA-BN within a population of healthcare personnel in Tshuapa province, DRC where clade I mpox occurs regularly (Clinicaltrials.gov registry number NCT02977715).^9^ This is this first evaluation of safety of MVA-BN in a clade I endemic country where MPXV circulates naturally.

## Methods and Materials

### Study design and Enrollment

The Centers for Disease Control and Prevention (CDC), DRC Ministry of Health (MOH), and Kinshasa School of Public Health (KSPH) implemented an open-label prospective cohort study of MVA-BN in healthcare personnel at risk of mpox. The study design has been described previously and is enrolled in clinicaltrials.gov (NCT02977715).^1^ Participants were followed on days 0, 14 (subset of total participants), 28, 42, 180, 365, 545, and 730. MVA-BN was administered subcutaneously as a 2-dose series on days 0 and 28 via either a liquid or lyophilized formulation; no one received a heterologous series. The liquid formulation was administered to participants from 4 health zones in Tshuapa Province and Kinshasa in 2017; and the lyophilized formulation was administered to participants in two health zones in Tshuapa Province in 2019 (Figure 1). Participants were excluded for any of the following reasons: positive pregnancy test on the day of vaccination; previous anaphylactic reaction to a vaccine, acute illness (e.g., temperature ≥37.2 °C, self-declaration, clinician discretion); other vaccine administration in the past 28 days; involvement in a research study in the last 4 weeks; and, presence of any conditions that may have placed the participant at an unacceptable risk of injury or rendered the participant unable to meet the requirements of the protocol (e.g., lack of mental capacity to consent, inability to complete follow up).

**Figure 1:**
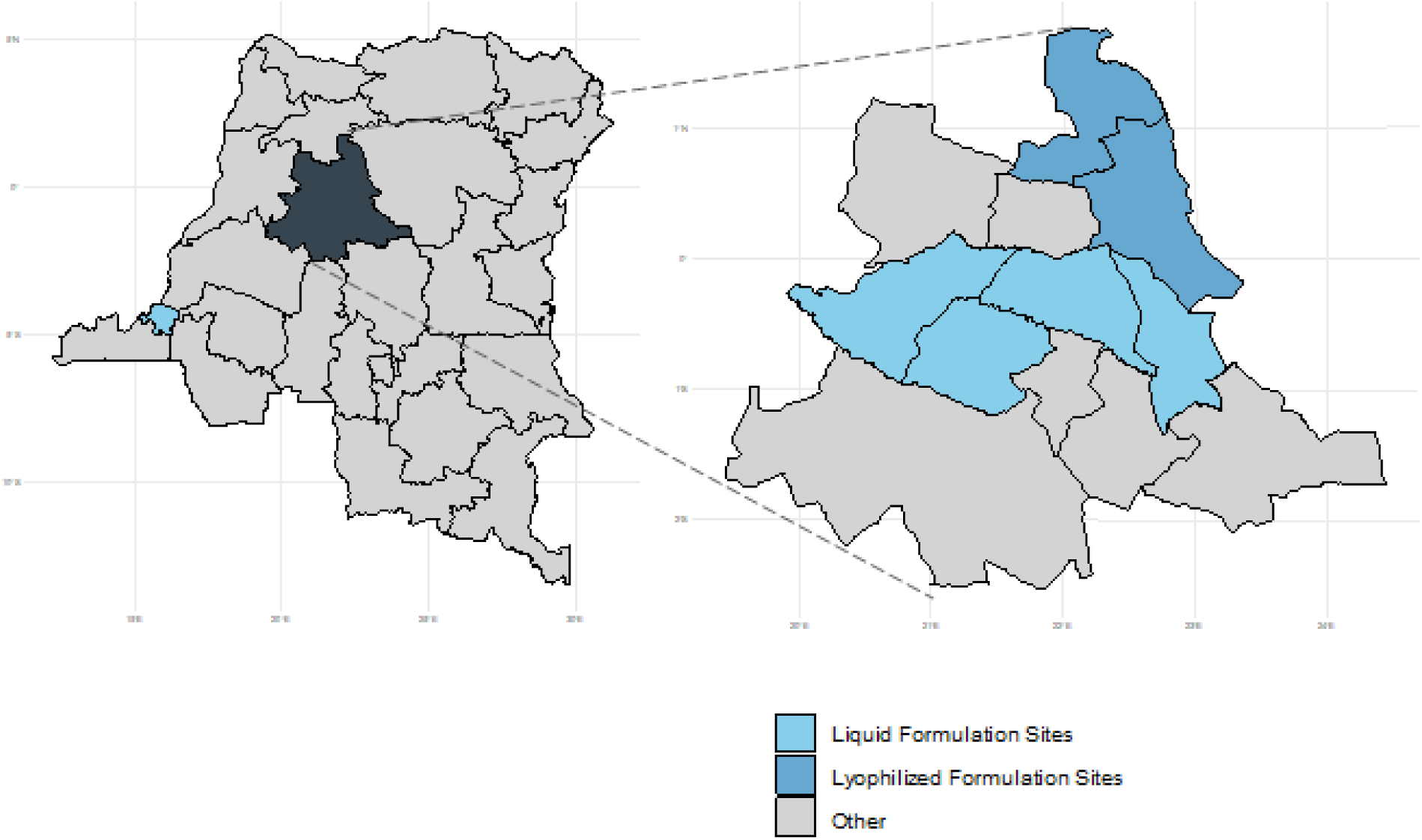
Locations of study sites. The figure represents the provinces of the Democratic Republic of the Congo. Highlighted on the left are Kinshasa and Tshuapa, the primary provinces of the study. The health zones within Tshuapa province (right) and the whole province of Kinshasa (light blue on left map) are colored based on where each formulation was used.

All female study participants of potential childbearing age (<60 years) had a pregnancy test administered upon initial enrollment prior to the first and, prior to second dose administration, were advised to avoid becoming pregnant for 28 days after vaccination. Participants who reported a new pregnancy or tested positive upon a rapid pregnancy test after receipt of the first vaccine dose did not receive the second vaccine dose but were followed for all subsequent study activities.

MVA-BN is manufactured by Bavarian Nordic. The vaccine comes in single dose vials with 1X10^8^ [TCID_50_] per 0.5 mL. Liquid and lyophilized formulations were used in 2017 and 2019, respectively, and the vaccine was stored, transported, and administered per manufacturer recommendations. Prior to use, the liquid formulation was thawed to room temperature, swirled for 30 seconds, and visually inspected for foreign particulate matter. The lyophilized formulation was brought to room temperature, reconstituted per manufacturer instructions, and visually inspected for foreign particulate matter. Vaccine was administered subcutaneously in the upper arm as a 2-dose series on days 0 and 28.

The protocol received human subjects’ ethical and regulatory approvals from the CDC^§^ and KSPH Institutional Review Boards and was authorized by the U.S. Food and Drug Administration. This study is registered in ClinicalTrials.gov under identifier NCT02977715.

### Safety Data collection

Data were collected from five forms: 1) enrollment; 2) self-report of medical history; 3) visit documentation; 4) serious adverse event reporting form completed by study staff; and 5) a 7-day adverse event diary completed by the participant. The initial enrollment, self-report of medical history and visit documentation forms recorded basic demographics, medical history, vaccine administration site, lot number, and dose data. Study staff observed participants for 30 minutes after vaccination for adverse events. Nineteen local and systemic reactions were solicited and recorded on the visit documentation form as either present or absent both prior to vaccination and 30 minutes after vaccine administration. Participants were provided with an adverse event diary containing six local and 13 systemic adverse reactions to monitor for the 7 days after vaccination. Participants were given a thermometer to measure temperature and instructed to complete the form each day. Participants were asked to record if their symptoms on each day limited daily activity or required medical intervention. Although cardiac events were of particular interest, the area where this study was conducted did not allow us to perform thorough cardiac evaluations (e.g., EKG, cardiac biomarkers). Therefore, assessment of myopericarditis was limited to reporting of chest pain symptoms in the symptom diary.

An adverse event (AE) was defined as any untoward medical occurrence associated with the use of MVA-BN, regardless of causal association with MVA-BN administration. Any AE or suspected AE was considered serious if it resulted in any of the following outcomes: death; life-threatening injury or illness; inpatient hospitalization or prolongation of existing hospitalization; persistent or significant incapacity or a substantial disruption of the ability to conduct normal life functions; congenital abnormalities or birth defects in the offspring of the study participant; or other important medical events that may jeopardize the participant or may require intervention to prevent one of the outcomes above. AEs were graded as 1- mild, 2-moderate, or 3-serious (Supplemental Appendix S0). Any grade 3 (serious AEs) prompted investigation by a local physician and completion of the serious AE reporting form. The physician submitted the form to the principal investigators and safety monitors. A minimum of two physician safety monitors (including board-certified physicians in the United States and in DRC) independently reviewed the report and assessed the relationship between MVA-BN and the occurrence of the event.

Data were collected on paper-based study forms and entered into a secure Microsoft Access database. Any conflicts in data entry were resolved through discussion among data entry managers and study personnel.

### Safety Data Analysis

RStudio version 4.0.3 was used to perform statistical analysis. All participants born in 1980 and after were considered vaccinia-naïve and those born before 1980 were conservatively considered previously immunized given routine childhood vaccination coverage with an orthopoxvirus vaccine was very high leading up to smallpox eradication in 1980. Demographic data were analyzed overall and compared by formulation. Adverse event data were analyzed and compared by formulation (overall for participants and stratified by dose) and by vaccination status stratified by formulation. Missing values were excluded in percentage calculations. Percentages were compared using chi-squared or Fisher’s Exact test or by calculating odds ratios and 95% confidence intervals. P-values were considered significant at <0.05.

## Results

### Participant characteristics

A total of 1600 participants were enrolled and received at least one dose of vaccine; 1000 received the liquid formulation and 600 received the lyophilized formulation. Of those administered vaccination, 1591 (99.4%) had forms returned and data available for analysis: 999 (99.9%) from the liquid formulation and 592 (98.7%) from the lyophilized formulation (Table 1). The median participant age was 45 (range 18– 84) years, 68% were male, and 74% of participants considered previously vaccinated. The second vaccine dose was given to 973 (97.4%) and 570 (96.3%) participants with the liquid and lyophilized formulations, respectively. Of the 48 (3.0%) participants who did not receive the second dose, 4 (8.3%) were due to pregnancy, 8 (16.7%) due to recent tetanus vaccination, 12 (25%) due to being absent or unenrolled, and the remaining 24 (50%) were lost to follow up. Three (0.2%) additional participants were lost to follow up after the second vaccination dose.

**Table 1.**
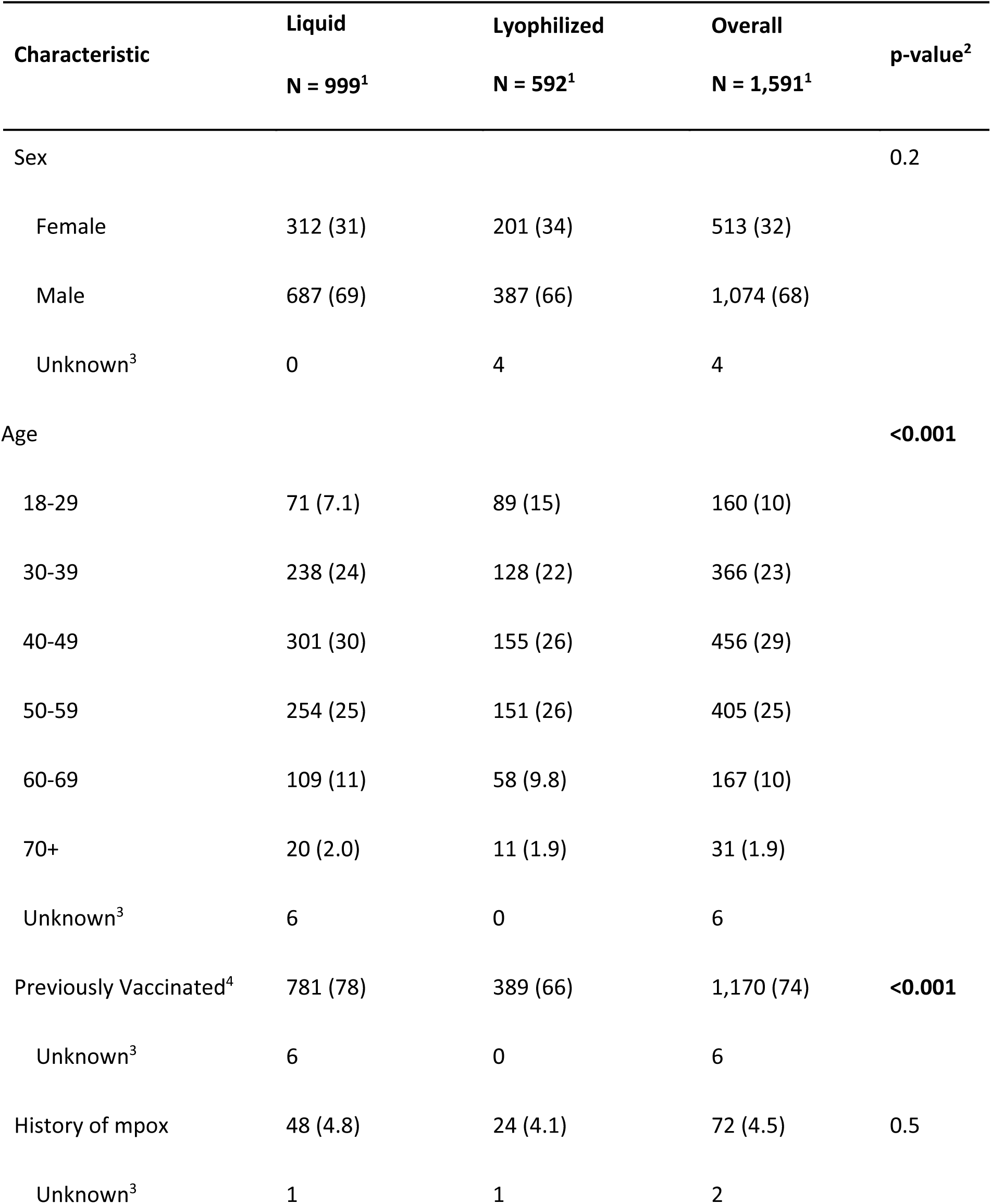
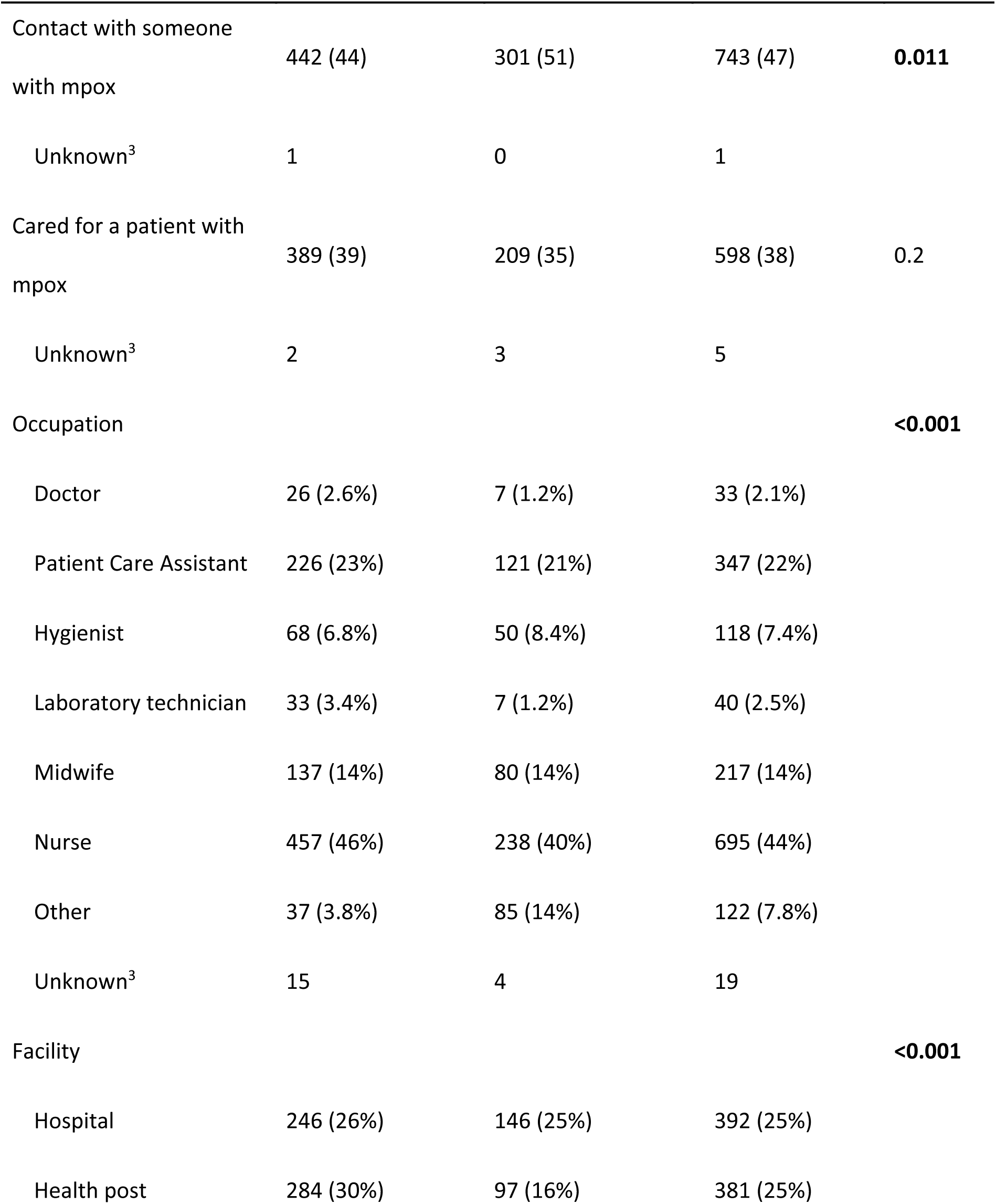
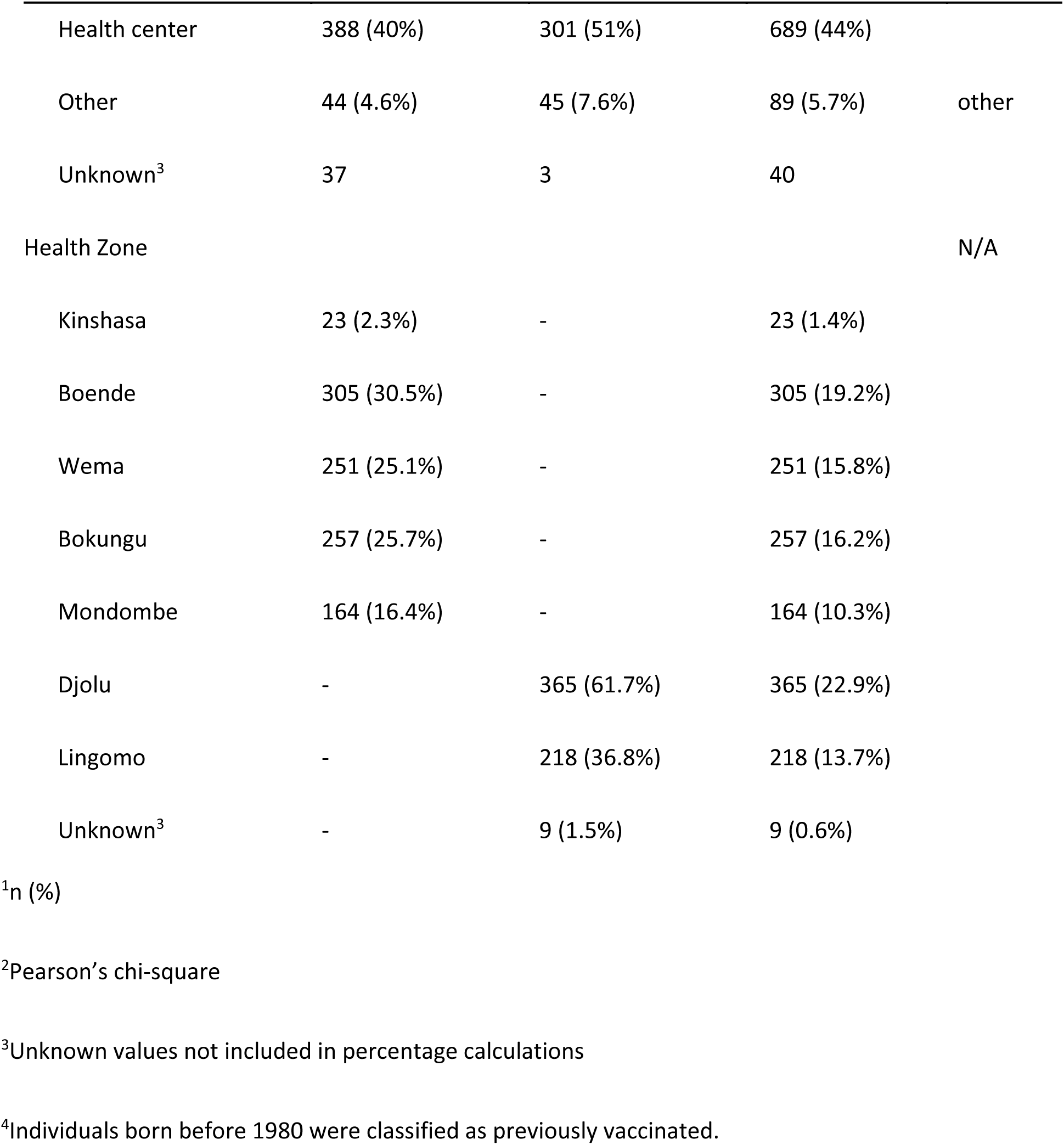
Demographic information of study participants.

### Safety analysis

Only four (0.2%) and one (<0.1%) of participants who received the liquid and lyophilized formulations, respectively, had any symptoms within 30 minutes of vaccine administration (Table S1). Within seven days after vaccination, participants noted similar proportions of local injection site reactions (36% and 36%) between the liquid and lyophilized formulations, respectively, but a lower proportion of systemic symptoms with the liquid (34%) compared to lyophilized (41%) formulation (Table 2). The most common injection site AEs reported included pain (24%), pruritus (13%), and edema (12%) for the liquid formulation and pain (20%), tenderness (13%), and pruritus (8.9%) for the lyophilized formulation. The most common systemic symptoms were headache (13%), fatigue (7.7%), and fever (7.5%) for the liquid formulation and headache (14%), fever (12%), and myalgia (10%) for the lyophilized formulation. Participants who received the liquid formulation had higher odds ratios for some injection site symptoms, including induration (OR 1.93 95% CI 1.26-3.04), edema (OR 1.84 95% CI 1.26-2.73), and pruritus (OR 1.54 95% CI 1.09-2.21). Participants who received the liquid formulation had lower odds ratio of any systemic symptoms (OR 0.73 95% CI 0.59-0.91), and of fever (OR 0.60 95% CI 0.42-0.86) and chills (OR 0.47 95% CI 0.30-0.74) specifically, compared to the lyophilized formulation.

**Table 2.**
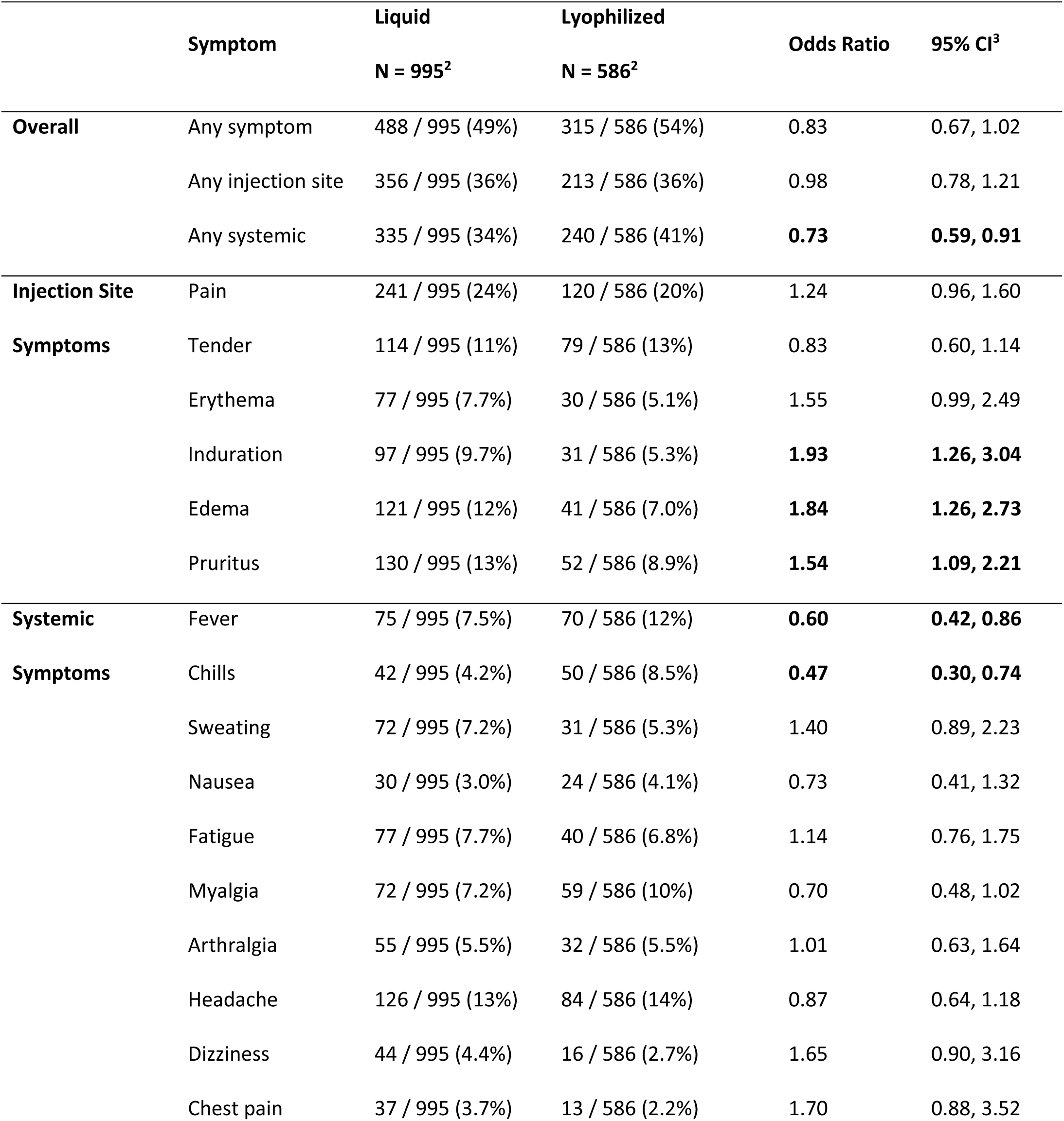
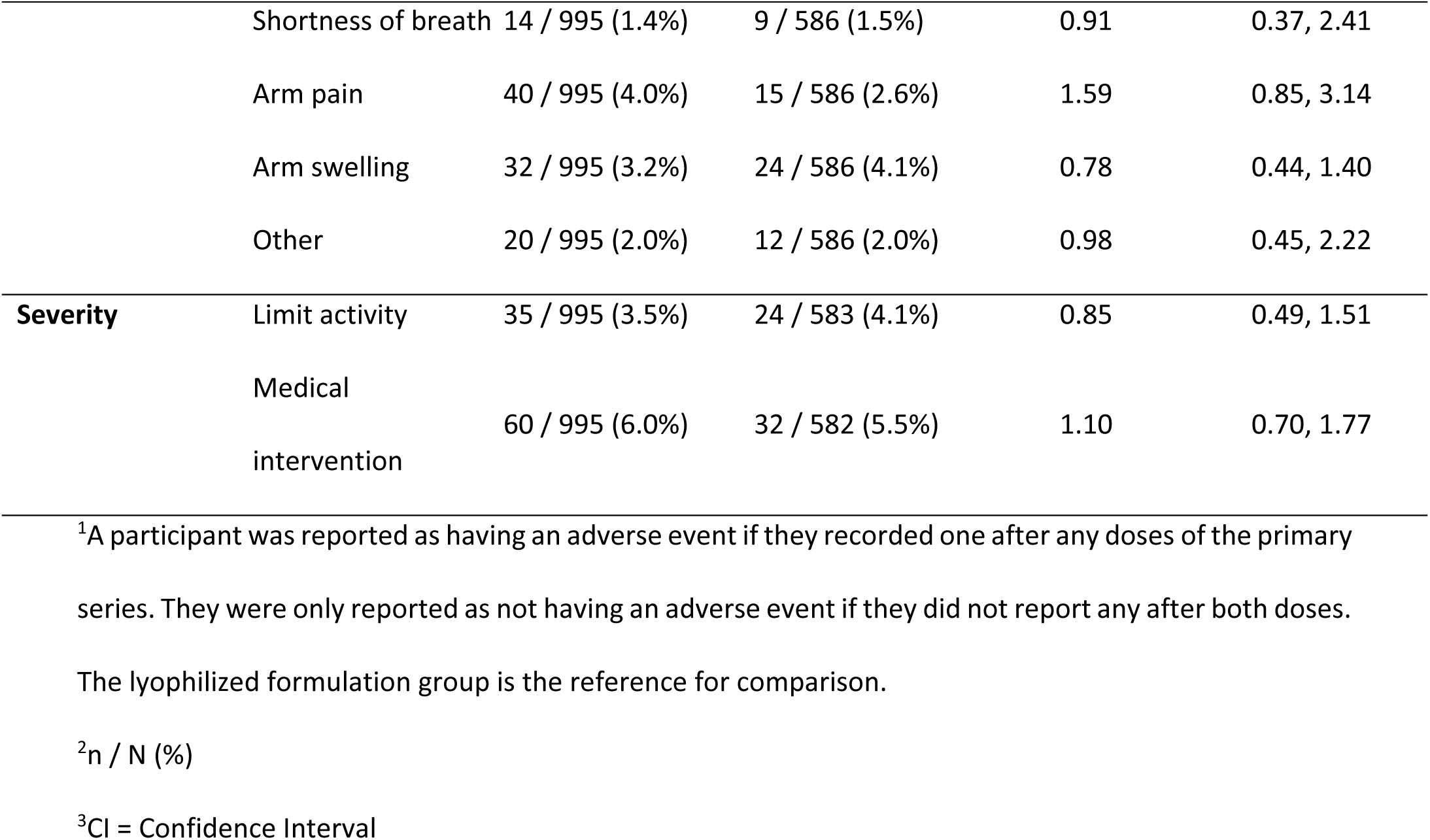
Number of participants that reported adverse events in the 7 days following vaccination by formulation^1^.

|  | Symptom | Liquid<br>N = 995<sup>2</sup> | Lyophilized<br>N = 586<sup>2</sup> | Odds Ratio | 95% CI<sup>3</sup> |
| --- | --- | --- | --- | --- | --- |
| **Overall** | Any symptom | 488 / 995 (49%) | 315 / 586 (54%) | 0.83 | 0.67, 1.02 |
|  | Any injection site | 356 / 995 (36%) | 213 / 586 (36%) | 0.98 | 0.78, 1.21 |
|  | Any systemic | 335 / 995 (34%) | 240 / 586 (41%) | **0.73** | **0.59, 0.91** |
| **Injection Site** | Pain | 241 / 995 (24%) | 120 / 586 (20%) | 1.24 | 0.96, 1.60 |
| **Symptoms** | Tender | 114 / 995 (11%) | 79 / 586 (13%) | 0.83 | 0.60, 1.14 |
|  | Erythema | 77 / 995 (7.7%) | 30 / 586 (5.1%) | 1.55 | 0.99, 2.49 |
|  | Induration | 97 / 995 (9.7%) | 31 / 586 (5.3%) | **1.93** | **1.26, 3.04** |
|  | Edema | 121 / 995 (12%) | 41 / 586 (7.0%) | **1.84** | **1.26, 2.73** |
|  | Pruritus | 130 / 995 (13%) | 52 / 586 (8.9%) | **1.54** | **1.09, 2.21** |
| **Systemic** | Fever | 75 / 995 (7.5%) | 70 / 586 (12%) | **0.60** | **0.42, 0.86** |
| **Symptoms** | Chills | 42 / 995 (4.2%) | 50 / 586 (8.5%) | **0.47** | **0.30, 0.74** |
|  | Sweating | 72 / 995 (7.2%) | 31 / 586 (5.3%) | 1.40 | 0.89, 2.23 |
|  | Nausea | 30 / 995 (3.0%) | 24 / 586 (4.1%) | 0.73 | 0.41, 1.32 |
|  | Fatigue | 77 / 995 (7.7%) | 40 / 586 (6.8%) | 1.14 | 0.76, 1.75 |
|  | Myalgia | 72 / 995 (7.2%) | 59 / 586 (10%) | 0.70 | 0.48, 1.02 |
|  | Arthralgia | 55 / 995 (5.5%) | 32 / 586 (5.5%) | 1.01 | 0.63, 1.64 |
|  | Headache | 126 / 995 (13%) | 84 / 586 (14%) | 0.87 | 0.64, 1.18 |
|  | Dizziness | 44 / 995 (4.4%) | 16 / 586 (2.7%) | 1.65 | 0.90, 3.16 |
|  | Chest pain | 37 / 995 (3.7%) | 13 / 586 (2.2%) | 1.70 | 0.88, 3.52 |
|  | Shortness of breath | 14 / 995 (1.4%) | 9 / 586 (1.5%) | 0.91 | 0.37, 2.41 |
|  | Arm pain | 40 / 995 (4.0%) | 15 / 586 (2.6%) | 1.59 | 0.85, 3.14 |
|  | Arm swelling | 32 / 995 (3.2%) | 24 / 586 (4.1%) | 0.78 | 0.44, 1.40 |
|  | Other | 20 / 995 (2.0%) | 12 / 586 (2.0%) | 0.98 | 0.45, 2.22 |
| **Severity** | Limit activity | 35 / 995 (3.5%) | 24 / 583 (4.1%) | 0.85 | 0.49, 1.51 |
|  | Medical<br>intervention | 60 / 995 (6.0%) | 32 / 582 (5.5%) | 1.10 | 0.70, 1.77 |

<sup>1</sup>A participant was reported as having an adverse event if they recorded one after any doses of the primary
series. They were only reported as not having an adverse event if they did not report any after both doses.
The lyophilized formulation group is the reference for comparison.
<sup>2</sup>n / N (%)
<sup>3</sup>CI = Confidence Interval

Reported AEs were more frequent following dose one compared to dose two for the liquid (43% and 23%, respectively) and lyophilized (47% and 17%, respectively) vaccine products (Table 3). The liquid formulation had a higher frequency of some local injection site reactions than the lyophilized formulation. For dose 1 only, this included pruritus (9.8% vs. 5.9%, p=0.007). For dose 2 only, this included tenderness (5.3% vs. 3.0%, p=0.042) and erythema (3.6% vs. 0.9%, p=0.002). For both dose 1 and dose 2, this included induration (7.4% vs. 4.3%, p=0.014 for dose 1 and 4.3% vs. 1.7%, p=0.007 for dose 2) and edema (9.0 vs. 5.2%, p=0.006 for dose 1 and 5.1% vs. 2.3%, p=0.008 for dose 2). The liquid formulation had a lower frequency for some systemic reactions compared to the lyophilized formulation. These included fever (4.6% vs 7.2%, p=0.026 for dose 1), chills (1.7% vs 7.1%, p<0.001 for dose 1), and myalgia (5.1% vs 9.0%, p=0.003 for dose 1). However, arm pain was reported at a higher frequency for the liquid formulation compared to the lyophilized formulation for dose 2 (1.8% vs 0.6%, p=0.049).

**Table 3.**
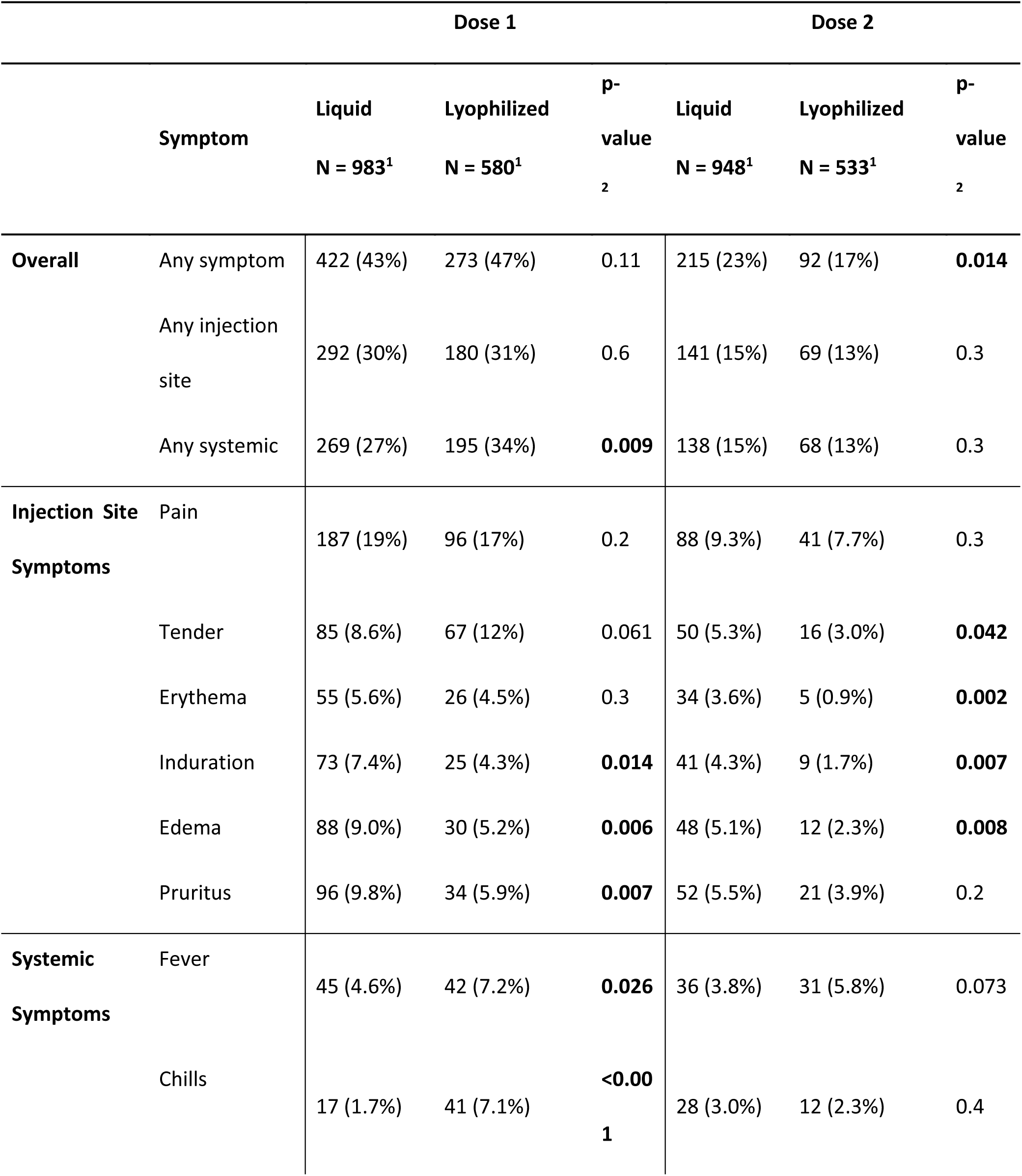
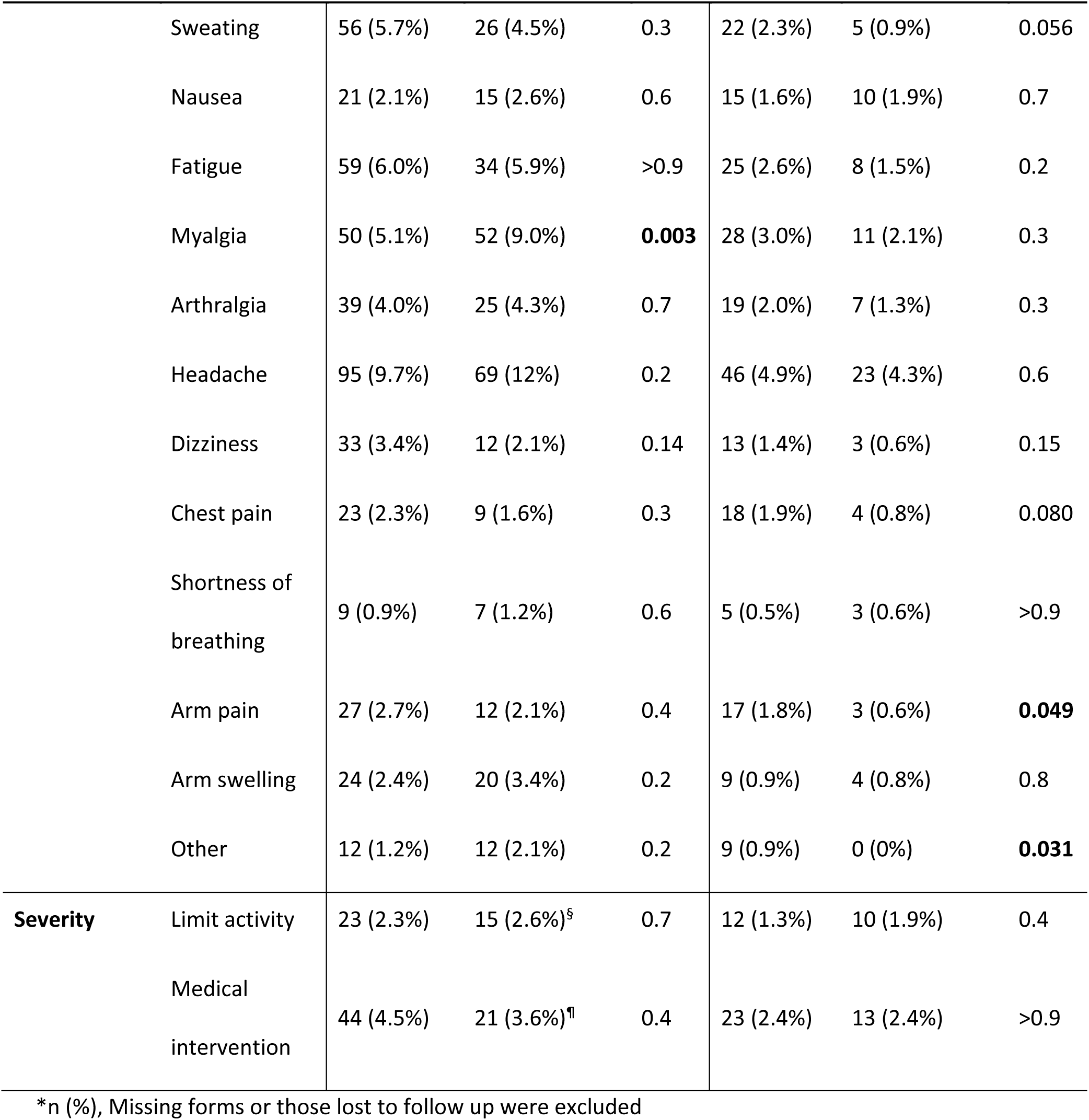
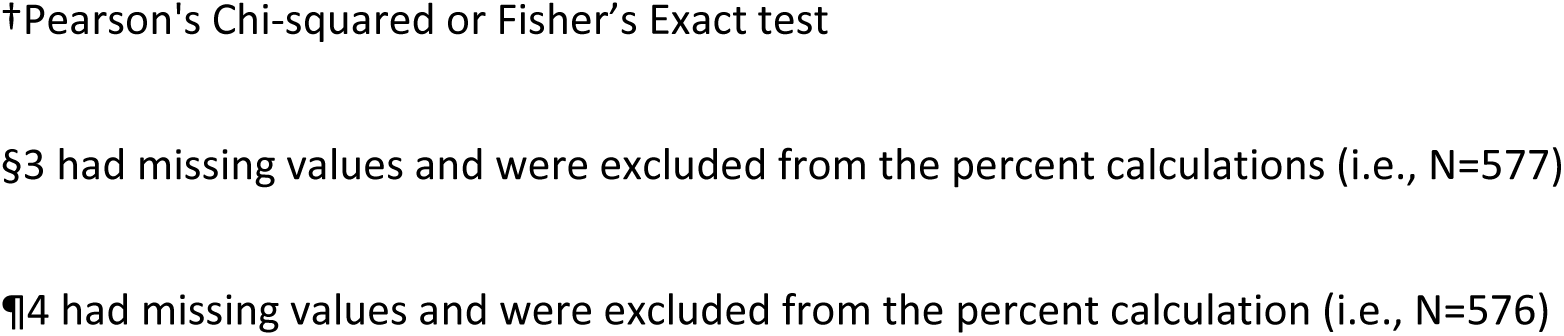
Frequency of reported adverse events in the 7 days following vaccination with MVA-BN by dose and formulation.

|  |  | Dose 1 |  |  | Dose 2 |  |  |
| --- | --- | --- | --- | --- | --- | --- | --- |
|  | Symptom | Liquid | Lyophilized | p- | Liquid | Lyophilized | p- |
|  |  | N = 983<sup>1</sup> | N = 580<sup>1</sup> | value<br>2 | N = 948<sup>1</sup> | N = 533<sup>1</sup> | value<br>2 |
| Overall | Any symptom | 422 (43%) | 273 (47%) | 0.11 | 215 (23%) | 92 (17%) | **0.014** |
|  | Any injection site | 292 (30%) | 180 (31%) | 0.6 | 141 (15%) | 69 (13%) | 0.3 |
|  | Any systemic | 269 (27%) | 195 (34%) | **0.009** | 138 (15%) | 68 (13%) | 0.3 |
| Injection Site Symptoms | Pain | 187 (19%) | 96 (17%) | 0.2 | 88 (9.3%) | 41 (7.7%) | 0.3 |
|  | Tender | 85 (8.6%) | 67 (12%) | 0.061 | 50 (5.3%) | 16 (3.0%) | **0.042** |
|  | Erythema | 55 (5.6%) | 26 (4.5%) | 0.3 | 34 (3.6%) | 5 (0.9%) | **0.002** |
|  | Induration | 73 (7.4%) | 25 (4.3%) | **0.014** | 41 (4.3%) | 9 (1.7%) | **0.007** |
|  | Edema | 88 (9.0%) | 30 (5.2%) | **0.006** | 48 (5.1%) | 12 (2.3%) | **0.008** |
|  | Pruritus | 96 (9.8%) | 34 (5.9%) | **0.007** | 52 (5.5%) | 21 (3.9%) | 0.2 |
| Systemic Symptoms | Fever | 45 (4.6%) | 42 (7.2%) | **0.026** | 36 (3.8%) | 31 (5.8%) | 0.073 |
|  | Chills | 17 (1.7%) | 41 (7.1%) | **<0.001** | 28 (3.0%) | 12 (2.3%) | 0.4 |
| Symptom |  | Liquid | Lyophilized | p- | Liquid | Lyophilized | p- |
|  |  | N = 983<sup>1</sup> | N = 580<sup>1</sup> | value | N = 948<sup>1</sup> | N = 533<sup>1</sup> | value |
|  |  |  |  | <sub>2</sub> |  |  | <sub>2</sub> |
|  | Sweating | 56 (5.7%) | 26 (4.5%) | 0.3 | 22 (2.3%) | 5 (0.9%) | 0.056 |
|  | Nausea | 21 (2.1%) | 15 (2.6%) | 0.6 | 15 (1.6%) | 10 (1.9%) | 0.7 |
|  | Fatigue | 59 (6.0%) | 34 (5.9%) | >0.9 | 25 (2.6%) | 8 (1.5%) | 0.2 |
|  | Myalgia | 50 (5.1%) | 52 (9.0%) | **0.003** | 28 (3.0%) | 11 (2.1%) | 0.3 |
|  | Arthralgia | 39 (4.0%) | 25 (4.3%) | 0.7 | 19 (2.0%) | 7 (1.3%) | 0.3 |
|  | Headache | 95 (9.7%) | 69 (12%) | 0.2 | 46 (4.9%) | 23 (4.3%) | 0.6 |
|  | Dizziness | 33 (3.4%) | 12 (2.1%) | 0.14 | 13 (1.4%) | 3 (0.6%) | 0.15 |
|  | Chest pain | 23 (2.3%) | 9 (1.6%) | 0.3 | 18 (1.9%) | 4 (0.8%) | 0.080 |
|  | Shortness of breathing | 9 (0.9%) | 7 (1.2%) | 0.6 | 5 (0.5%) | 3 (0.6%) | >0.9 |
|  | Arm pain | 27 (2.7%) | 12 (2.1%) | 0.4 | 17 (1.8%) | 3 (0.6%) | **0.049** |
|  | Arm swelling | 24 (2.4%) | 20 (3.4%) | 0.2 | 9 (0.9%) | 4 (0.8%) | 0.8 |
|  | Other | 12 (1.2%) | 12 (2.1%) | 0.2 | 9 (0.9%) | 0 (0%) | **0.031** |
| **Severity** | Limit activity | 23 (2.3%) | 15 (2.6%)<sup>§</sup> | 0.7 | 12 (1.3%) | 10 (1.9%) | 0.4 |
|  | Medical intervention | 44 (4.5%) | 21 (3.6%)<sup>¶</sup> | 0.4 | 23 (2.4%) | 13 (2.4%) | >0.9 |

\*n (%), Missing forms or those lost to follow up were excluded
†Pearson's Chi-squared or Fisher's Exact test
§3 had missing values and were excluded from the percent calculations (i.e., N=577)
¶4 had missing values and were excluded from the percent calculation (i.e., N=576)

Most AEs were mild with either formulation. Overall, less than 5% of participants had AEs that limited activity, and less than 7% of participants had AEs requiring medical intervention, all considered a grade-2 (moderate) AE. There was a statistically significantly higher rate of medical intervention needed for participants who received dose one of the liquid formulation compared to dose two (4.5% vs 2.4%, p=0.015) however, this finding was not significant with the lyophilized formulation (3.6% vs 2.4%, p=0.091). Chest pain was reported in 3.7% vs 2.2% of participants for liquid and lyophilized formulations, respectively.

Participants who were considered previously vaccinated had a lower odds ratio of experiencing at least one symptom, regardless of formulation (OR 0.64, 95% CI 0.47-0.87 for liquid, OR 0.63, 95% CI 0.44-0.91 for lyophilized) (Table 4).

**Table 4:** Frequencies and Odds Ratios of adverse events by vaccination status^1^ stratified by formulation in the 7 days following vaccination.

| Liquid Formulation |  |  |  |  | Lyophilized Formulation |  |  |  |
| --- | --- | --- | --- | --- | --- | --- | --- | --- |
| Characteristic | Previously<br>Vaccinated<br>N = 773<sup>2</sup> | Vaccinia Naïve<br>N = 222<sup>2</sup> | Odds<br>Ratio | 95% CI<sup>3</sup> | Previously<br>Vaccinated<br>N = 385<sup>2</sup> | Vaccinia Naïve<br>N = 201<sup>2</sup> | Odds<br>Ratio | 95% CI<sup>3</sup> |
| **Overall** |  |  |  |  |  |  |  |  |
| Any Symptom | 360 / 773 (47%) | 128 / 222 (58%) | **0.64** | **0.47, 0.87** | 192 / 385 (50%) | 123 / 201 (61%) | **0.63** | **0.44, 0.91** |
| Any Injection<br>Symptom | 254 / 773 (33%) | 102 / 222 (46%) | **0.58** | **0.42, 0.79** | 123 / 385 (32%) | 90 / 201 (45%) | **0.58** | **0.40, 0.84** |
| Any Systemic<br>Symptom | 246 / 773 (32%) | 89 / 222 (40%) | **0.70** | **0.51, 0.96** | 148 / 385 (38%) | 92 / 201 (46%) | 0.74 | 0.52, 1.06 |
| **Injection Site Symptoms** |  |  |  |  |  |  |  |  |
| Pain | 164 / 773 (21%) | 77 / 222 (35%) | **0.51** | **0.36, 0.71** | 72 / 385 (19%) | 48 / 201 (24%) | 0.73 | 0.48, 1.14 |
| Tender | 75 / 773 (9.7%) | 39 / 222 (18%) | **0.50** | **0.33, 0.79** | 46 / 385 (12%) | 33 / 201 (16%) | 0.69 | 0.42, 1.16 |
| Erythema | 53 / 773 (6.9%) | 24 / 222 (11%) | 0.61 | 0.36, 1.06 | 17 / 385 (4.4%) | 13 / 201 (6.5%) | 0.67 | 0.30, 1.53 |
| Induration | 66 / 773 (8.5%) | 31 / 222 (14%) | **0.58** | **0.36, 0.94** | 14 / 385 (3.6%) | 17 / 201 (8.5%) | **0.41** | **0.18, 0.90** |
| Edema | 87 / 773 (11%) | 34 / 222 (15%) | 0.70 | 0.45, 1.11 | 19 / 385 (4.9%) | 22 / 201 (11%) | **0.42** | **0.21, 0.84** |
| Characteristic | Previously<br>Vaccinated<br>N = 773<sup>2</sup> | Vaccinia Naïve<br>N = 222<sup>2</sup> | Odds<br>Ratio | 95% CI<sup>3</sup> | Previously<br>Vaccinated<br>N = 385<sup>2</sup> | Vaccinia Naïve<br>N = 201<sup>2</sup> | Odds<br>Ratio | 95% CI<sup>3</sup> |
| Pruritus | 97 / 773 (13%) | 33 / 222 (15%) | 0.82 | 0.53, 1.30 | 32 / 385 (8.3%) | 20 / 201 (10.0%) | 0.82 | 0.44, 1.56 |
| **Systemic Symptoms** |  |  |  |  |  |  |  |  |
| Fever | 50 / 773 (6.5%) | 25 / 222 (11%) | **0.55** | **0.32, 0.94** | 51 / 385 (13%) | 19 / 201 (9.5%) | 1.46 | 0.82, 2.71 |
| Chills | 22 / 773 (2.8%) | 20 / 222 (9.0%) | **0.30** | **0.15, 0.58** | 32 / 385 (8.3%) | 18 / 201 (9.0%) | 0.92 | 0.49, 1.79 |
| Sweating | 52 / 773 (6.7%) | 20 / 222 (9.0%) | 0.73 | 0.42, 1.32 | 17 / 385 (4.4%) | 14 / 201 (7.0%) | 0.62 | 0.28, 1.39 |
| Nausea | 19 / 773 (2.5%) | 11 / 222 (5.0%) | 0.48 | 0.21, 1.14 | 12 / 385 (3.1%) | 12 / 201 (6.0%) | 0.51 | 0.20, 1.26 |
| Fatigue | 49 / 773 (6.3%) | 28 / 222 (13%) | **0.47** | **0.28, 0.80** | 24 / 385 (6.2%) | 16 / 201 (8.0%) | 0.77 | 0.38, 1.59 |
| Myalgia | 49 / 773 (6.3%) | 23 / 222 (10%) | 0.59 | 0.34, 1.03 | 30 / 385 (7.8%) | 29 / 201 (14%) | **0.50** | **0.28, 0.90** |
| Arthralgia | 37 / 773 (4.8%) | 18 / 222 (8.1%) | 0.57 | 0.31, 1.09 | 21 / 385 (5.5%) | 11 / 201 (5.5%) | 1.00 | 0.45, 2.34 |
| Headache | 86 / 773 (11%) | 40 / 222 (18%) | **0.57** | **0.37, 0.88** | 47 / 385 (12%) | 37 / 201 (18%) | 0.62 | 0.38, 1.02 |
| Dizziness | 38 / 773 (4.9%) | 6 / 222 (2.7%) | 1.86 | 0.77, 5.46 | 11 / 385 (2.9%) | 5 / 201 (2.5%) | 1.15 | 0.36, 4.29 |
| Chest pain | 24 / 773 (3.1%) | 13 / 222 (5.9%) | 0.52 | 0.25, 1.12 | 6 / 385 (1.6%) | 7 / 201 (3.5%) | 0.44 | 0.12, 1.55 |
| Shortness of breath | 12 / 773 (1.6%) | 2 / 222 (0.9%) | 1.73 | 0.38, 16.1 | 4 / 385 (1.0%) | 5 / 201 (2.5%) | 0.41 | 0.08, 1.94 |
| Characteristic | Previously<br>Vaccinated<br>N = 773<sup>2</sup> | Vaccinia Naïve<br>N = 222<sup>2</sup> | Odds<br>Ratio | 95% CI<sup>3</sup> | Previously<br>Vaccinated<br>N = 385<sup>2</sup> | Vaccinia Naïve<br>N = 201<sup>2</sup> | Odds<br>Ratio | 95% CI<sup>3</sup> |
| Arm pain | 24 / 773 (3.1%) | 16 / 222 (7.2%) | **0.41** | **0.21, 0.85** | 8 / 385 (2.1%) | 7 / 201 (3.5%) | 0.59 | 0.18, 1.94 |
| Arm swelling | 23 / 773 (3.0%) | 9 / 222 (4.1%) | 0.73 | 0.32, 1.81 | 7 / 385 (1.8%) | 17 / 201 (8.5%) | **0.20** | **0.07, 0.52** |
| Other | 14 / 773 (1.8%) | 6 / 222 (2.7%) | 0.66 | 0.24, 2.14 | 6 / 385 (1.6%) | 6 / 201 (3.0%) | 0.52 | 0.14, 1.95 |
| **Severity** |  |  |  |  |  |  |  |  |
| Limit activity | 26 / 773 (3.4%) | 9 / 222 (4.1%) | 0.82 | 0.37, 2.03 | 14 / 384 (3.6%) | 10 / 199 (5.0%) | 0.72 | 0.29, 1.84 |
| Medical intervention | 40 / 773 (5.2%) | 20 / 222 (9.0%) | 0.55 | 0.31, 1.02 | 20 / 383 (5.2%) | 12 / 199 (6.0%) | 0.86 | 0.39, 1.97 |

<sup>1</sup>Previously Vaccinated defined as those born prior to 1980
<sup>2</sup>n / N (%)
<sup>3</sup>CI = Confidence Interval

Eighteen grade 3 (serious) AEs, including seventeen deaths (rate 5.1 deaths/1,000 person-year (95% CI 1.6-10.2)) and one stillbirth were reported and investigated during the two-year study period (Table S2). Study safety monitors determined that none of the serious AEs were related to MVA-BN administration.

### Pregnancies

Fourteen pregnancies occurred within 28 days of vaccine administration (Table S3). Thirteen (93%) resulted in healthy infants born at or near term (35-41 weeks) and one (7%) resulted in a stillbirth at 37 weeks gestation, reported as a serious AE, and determined by study safety monitors not to be related to MVA-BN administration.

## Discussion

This study provides data from the safety monitoring for the first field trial of MVA-BN vaccine in an endemic area of Africa. The large number of participants and duration of follow-up ensured comprehensive safety monitoring data. Among study participants, the AE frequency was 49% and 54% for liquid and lyophilized formulations, respectively. Most AEs were mild and did not require additional treatment or limit activity level. Safety monitoring did not identify any serious AEs related to vaccine administration.

Data from this study demonstrate lower rates of AEs compared to rates observed from published clinical trials. Thirty-six percent of participants experienced a local vaccination site reaction for both formulations and 34% and 41% of participants reported a systemic reaction with the liquid and lyophilized formulations, respectively. AE rates were higher with naïve participants (58% of the liquid and 61% of the lyophilized), compared to previously vaccinated participants (47% for liquid and 50% for lyophilized). In comparison to the largest (n=4005) phase III clinical trials in vaccination naïve participants, the overall rates of solicited local or systemic AEs were 88.6-90.6% in groups receiving MVA-BN.^10^ In another study with previously vaccinated participants, the rates of solicited AEs were slightly lower at 72.6-85.5% for local and 33.9-43.5%, for systemic AEs.^11^ AE rates reported 7 days after vaccination in a study from Australia were similar to this study at 44% for local and 23% for systemic AEs.^12^ However, in a study in Italy, AE rates were higher, 93% with local and 48% with systemic AEs.^13^

There are several possible reasons for the observed decreased AE rate in our study. There may be cultural differences between what would be considered an adverse reaction worth self-reporting. Additionally, many diaries were not completed daily and were completed at the subsequent study visit with the assistance of study staff, which may help with reporting but, also, could result in recall bias.

Most participants (98.6%) from this study population live and work in Tshuapa Province where MPXV is endemic.^14,15^ Although most of the population (74%) in this study were born before 1980 and were likely previously vaccinated for smallpox, many remain at risk due to the nature of their work; 47% reported having an exposure to someone with mpox previously and 38% reporting caring for someone with mpox in the past. Study participants were expected to have a relatively higher level of baseline exposure to OPXV then populations used during the clinical trials in Europe and the United States.^1^ Studies in the United States and in Europe have suggested that vaccinia naïve populations may experience higher rates of AEs.^10,13,16^ We similarly found that vaccinia naïve (i.e., previously unvaccinated) individuals had 1.56-1.58 greater odds ratio of experiencing at least one AE.

Seventeen deaths were identified during the study period, the US and DRC-based study physician safety monitors determined these deaths were unrelated to vaccine administration. The annual crude death rate in DRC is 9.9 deaths/1,000 persons based on 2017 estimates.^17^ By comparison, the annual death rate among the study participants was lower than the overall country estimates at 5.1 deaths/1,000 persons (95% CI 1.6-10.2). The number of deaths seen in participants is not unexpected for this population.

The results of this study need to be interpreted considering some limitations. The symptoms in the AE diary were self-reported and were not clinically evaluated or measured individually in terms of severity. Each symptom was asked about separately, then participants were collectively asked about limitations in activity and if medical interventions were required. There may be some residual confounding associated with adverse event rates given most patients in the previously vaccinated group were older. Secondly, access to healthcare and treatment in Tshuapa is limited. Study physicians were limited by the quality of health records and access to imaging and laboratory diagnostics to determine diagnoses when investigating SAEs. Thirdly, there were a small number of pregnant people in this study and results should be interpreted cautiously. Finally, no active surveillance for myopericarditis was conducted, such as, ECG or serial troponin levels. Therefore, participants experiencing chest pain could not be thoroughly evaluated for myopericarditis.

## Conclusions

This study adds to the growing body of literature on the safety of MVA-BN in different populations. The data from the Congolese healthcare personnel cohort demonstrate good and similar safety outcomes for up to two years with both liquid and lyophilized vaccine product. The increased stability of the lyophilized formulation may help reach communities with storage and transportation challenges. Given the increase in clade I mpox and spread to surrounding countries, these data provide reassurance of the safety of this vaccine in endemic regions. As MVA-BN vaccination campaigns are launched in endemic and surrounding countries in late 2024, the data from this study supporting safety of MVA-BN in an African population are crucial to build vaccine confidence. However, continued pharmacovigilance is necessary to capture any AEs once larger populations are vaccinated.

^§^See 45 C.F.R. part 46; 21 C.F.R. part 5

## Data Availability

All data produced in the present work are contained in the manuscript

## Acknowledgements

The authors of this paper would like to thank colleagues from the US Centers for Disease Control and Prevention, the Kinshasa School of Public Health, the Congolese Ministry of Health and the University of Kinshasa for their contributions to this study.

