## Supplemental Appendix for "Safety of MVA-BN in Healthcare Personnel, Democratic Republic of the Congo"

### Section S0: Grading of adverse events

- Grade 1-Mild: Transient or mild discomforts (usually < 48 hours), no or minimal medical intervention/ therapy required, hospitalization not necessary (non-prescription or single-use prescription therapy may be employed to relieve symptoms (e.g., aspirin for a headache)
- Grade 2-Moderate: Mild to moderate limitation in activity, some assistance may be needed; no or minimal intervention/ therapy required, hospitalization possible
- Grade 3-serious: Marked limitation in activity; medical intervention/therapy or hospitalization to prevent permanent disability, life-threatening condition, or death

### Table S1: Reported frequencies of symptoms prior to and 30 minutes after MVA-BN administration

| | **Liquid** | | **Lyophilized** | |
|---|---|---|---|---|
| **Characteristic** | **Before** N = 1,971[1] | **After** N = 1,971[1] | **Before** N = 1,173[1] | **After** N = 1,173[1] |
| Any Symptom | 45/1967 (2.3%) | 4/1959 (0.2%) | 6/1173 (0.5%) | 1/1173 (<0.1%) |
| Chills | 6/1970 (0.3%) | 1/1961 (<0.1%) | 1/1173 (<0.1%) | 0/1173 (0%) |
| Sweating | 1/1970 (<0.1%) | 0/1961 (0%) | 0/1173 (0%) | 0/1173 (0%) |
| Fever | 3/1969 (0.2%) | 0/1961 (0%) | 0/1173 (0%) | 0/1173 (0%) |
| Nausea | 2/1970 (0.1%) | 0/1961 (0%) | 0/1173 (0%) | 0/1173 (0%) |
| Fatigue | 6/1970 (0.3%) | 0/1961 (0%) | 1/1173 (<0.1%) | 0/1173 (0%) |
| Myalgia | 8/1970 (0.4%) | 0/1960 (0%) | 0/1173 (0%) | 0/1173 (0%) |
| Arthralgia | 6/1969 (0.3%) | 1/1961 (<0.1%) | 0/1173 (0%) | 0/1173 (0%) |
| Headache | 16/1970 (0.8%) | 1/1961 (<0.1%) | 3/1173 (0.3%) | 0/1173 (0%) |
| Dizziness | 6/1970 (0.3%) | 1/1959 (<0.1%) | 0/1173 (0%) | 1/1173 (<0.1%) |
| Chest pain | 8/1968 (0.4%) | 0/1961 (0%) | 0/1173 (0%) | 0/1173 (0%) |
| Shortness of breath | 4/1968 (0.2%) | 0/1961 (0%) | 0/1173 (0%) | 0/1173 (0%) |
| Arm pain | 4/1967 (0.2%) | 0/1961 (0%) | 0/1173 (0%) | 0/1173 (0%) |
| Arm swelling | 1/1961 (<0.1%) | 0/1960 (0%) | 1/1173 (<0.1%) | 0/1173 (0%) |

[1]Includes both first and second doses

### Table S2. Serious adverse event reports

| Participant number | Formulation | Sex | Age group | Event | Time to event from last vaccine dose | Cause of SAE per clinician review and report |
|---|---|---|---|---|---|---|
| 1 | Liquid | F | 60-69 | Death | 11 months | Head injury |
| 2 | Liquid | F | 30-39 | Death | 8 months | Acute hepatitis with severe anemia |
| 3 | Liquid | M | 40-49 | Death | 3 months | Not specified; patient presented with swelling and ulceration of the leg, headaches, dyspnea, fever; patient consulted with a traditional healer |
| 4 | Liquid | M | 50-59 | Death | 10 months | Not specified; the patient did not seek medical care and died at home after unquantified weight loss and prolonged fever |
| 5 | Liquid | M | 70+ | Death | 9 months | Acute gastroenteritis associated with severe malaria |
| 6 | Liquid | M | 70+ | Death | 4 months | Not specified; the patient had a diagnosis of hypertensive encephalopathy and malaria |
| 7 | Liquid | M | 30-39 | Death | 8 months | Alcohol intoxication |
| 8 | Liquid | M | 40-49 | Death | 2 months | Cerebral Vascular Accident;<br>Cryptococcus neoformans in a patient infected with HIV |
| 9 | Liquid | M | 50-59 | Death | 1 week | Alcoholic encephalopathy, hypoglycemia, lung disease |
| 10 | Liquid | M | 40-49 | Death | 1 year 4 months | Complications secondary to hepatic cirrhosis including hepatic encephalopathy, ascites, respiratory distress |
| 11 | Liquid | M | 40-49 | Death | 1 year 6 months | Opportunistic infections secondary to advanced HIV |
| 12 | Liquid | M | 30-39 | Death | 1 year 9 months | Liver cancer and hepatitis B complicated by ascites |

| | | | | | | |
|---|---|---|---|---|---|---|
| 13 | Liquid | F | | Still birth | 9 months | Received second dose of vaccine likely prior to or days near conception. Stillbirth at 37 weeks (weight 1000g), |
| 14 | Lyophilized | F | 40-49 | Death | 1 year 3 months | Cerebrovascular event in the context of advanced HIV |
| 15 | Lyophilized | M | 70+ | Death | 1 year 5 months | Hypertensive cardiomyopathy complicated by severe anemia (Hgb, 6 mg/dL) |
| 16 | Lyophilized | M | 40-49 | Death | 1 year 3 months | Right inguinal/scrotal hernia complicated by septic shock |
| 17 | Lyophilized | M | 30-39 | Death | 11 months | Tonic clonic movements followed by sudden cardiac arrest |
| 18 | Lyophilized | M | 70+ | Death | 1 year, 11 months | Septic shock secondary to necrotic ulcer from a machete wound >8 months prior |
